# Synthetic protein antigens for COVID-19 diagnostics

**DOI:** 10.1101/2021.02.23.21251934

**Authors:** Catherine H. Schein, Corri B. Levine, Susan L F McLellan, Surendra S. Negi, Werner Braun, Stephen C. Dreskin, Elizabeth S. Anaya, Jurgen Schmidt

**Affiliations:** Department of Biochemistry and Molecular Biology, The University of Texas Medical Branch, Galveston, TX; Institute for Human Infections and immunity (IHII), The University of Texas Medical Branch, Galveston, TX; Institute for Translational Sciences, The University of Texas Medical Branch, Galveston, TX; Department of Internal medicine – Infectious Diseases, The University of Texas Medical Branch, Galveston, TX; Sealy Center for Structural Biology and Molecular Biophysics, The University of Texas Medical Branch, Galveston, TX; Division of Allergy and Clinical Immunology, Department of Medicine, University of Colorado Denver, Aurora, CO. 80045; B-11 Bioenergy and Biome Sciences, Bioscience Division Los Alamos National Laboratory, Los Alamos, NM 87545

**Keywords:** COVID-19 variants, Structure based design, Receptor binding domain, S protein epitopes, ACE2 interaction

## Abstract

There is an urgent need for inexpensive, rapid and specific antigen-based assays to test for infection with SARS-CoV-2 and distinguish variants arising as the COVID-19 pandemic spreads. We have identified a small, synthetic protein (JS7), representing a region of maximum variability within the receptor binding domain (RBD), which binds antibodies in sera from nine patients with PCR-verified COVID-19 of varying severity. Antibodies binding to either JS7 or the SARS-CoV-2 recombinant RBD, as well as those that disrupt binding between a fragment of the ACE2 receptor and the RBD, are proportional to disease severity and clinical outcome. Binding to JS7 was inhibited by linear peptides from the RBD interface with ACE2. Variants of JS7, such as N501Y, can be quickly synthesized in a pure form in large quantities by automated methods. JS7 and related synthetic antigens can provide a basis for specific diagnostics for SARS-CoV-2 infections.

## Introduction

COVID-19, caused by a new coronavirus (SARS-CoV-2) ^5,6^ is closely related, in its sequence, structure ^8^, binding to the human ACE2 receptor ^10^ and epitopes recognized by neutralizing antibodies isolated from survivors ^1, 11^ of the SARS-CoV-1 (SARS) 2002-2003 outbreak ^12, 13^ and COVID-19 ^14^. In addition, the previously identified SARS-specific human antibody, CR3022, binds the receptor binding domain (RBD) of SARS-Cov-2 with nanomolar affinity ^16^. Practical tools to distinguish these viruses and especially the recently identified variants of SARS-CoV-2 ^15, 17^ are needed. However, the viruses diverge from each other in that the case fatality rate of the newer virus appears lower. SARS-CoV-2 is much more communicable via inhaled aerosols ^6,18^, perhaps due to insertion of a furin cleavage site around position 701^19^. SARS-CoV-2 has continued to mutate during its path through humanity, accumulating changes in this furin site and the RBD that may eventually affect its phenotype, immune sensitivity and resistance to therapies ^4,20-23^. Although both SARS viruses are quite distinct in sequence from the even deadlier β-CoV that causes MERS ^24^, variants or recombinants of these pathogens could result in a virus that combines the high transmissibility of SARS-CoV-2 with the high mortality of SARS-CoV-1 or MERS. In addition, while the origins of both SARS viruses are unclear, there is a high probability that similar viruses are circulating, re-combining and mutating in some animal reservoir. Cross-over of the human virus into other species could also lead to changes in its infectivity severity, and resistance to therapies^25, 26^.It is thus essential to have sensitive and specific diagnostic tools for the public health community to detect currently circulating variants. Outbreaks with mutated forms of SARS-CoV-2 are continuing to emerge ^15, 22, 27^. Many assays for determining prior COVID-19 infection are based on detecting antibodies in sera to recombinant versions of the surface (S) protein, which was previously identified as the binding site for neutralizing antibodies produced in response to SARS-CoV-1. At least one SARS-CoV-1 neutralizing monoclonal antibody ^28, 29^ also binds with less affinity to the S protein SARS-CoV-2 ^30^, which varies by about 20% throughout its length (Fig. S1). Fortunately, methods developed for expressing the SARS S protein could be adapted to express also that of SARS-Cov-2, and the RBD, despite the sequence and length (1255/1277 aa for SARS/SARS-CoV2) diversity^7,13^.However, the S proteins of future variants may not be as amenable to recombinant synthesis.

Recent advances allow chemical synthesis of small proteins in large amounts, supplying pure protein variants rapidly. We have thus concentrated on reducing the size of the protein needed to distinguish COVID-19 infections to the area of maximum sequence variation between SARS-CoV-1 and SARS-CoV-2 ^9,31-33^. This area also coincides with a region where the epitopes of many COVID-19 neutralizing monoclonal antibodies cluster.^34^ As we show here, synthetic proteins representing this region bind antibodies from sera of patients with COVID-19 infections of varying severity. Modifications of this protein can be the basis for detecting variants that may affect treatment protocols.

## Results and Discussion

We synthesized two proteins, of about 10 kD and 7 KD (JS10 and JS7, Fig. 1, see Supplementary Material for production and characterization of the latter). This area of the RBD structure, in complex with the ACE2 in a cryo-EM structure^35^, contains a region of antiparallel β-sheet (Fig. 1), but is otherwise flexibly structured with 4 tyrosines (Y453, Y489, Y449, Y505) mediating many contacts across the interface. The recent UK mutations, N501Y, could enhance this binding.The CD spectrum of JS7 (Fig. S2) suggests it forms the flexible structure and β-strands that characterize the experimental structure.

**Fig. 1.**
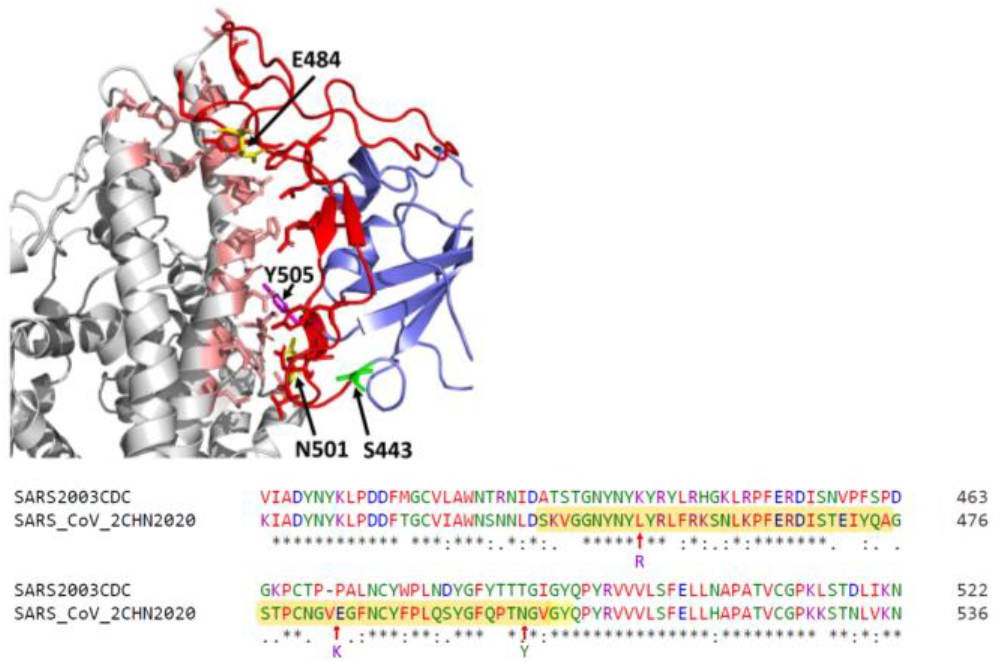
The JS7 synthetic protein (S443-Y505, blue line) represents the most variable region of SARS-CoV-2 S protein (relative to SARS-CoV-1 from 2003) which mediates its interaction with ACE2 Top) A cryo-EM structure (PDB entry 7KMB) of the complex shows how the JS7 segment (red) of the RBD (blue) lies at the intersection with the ACE2 cell receptor (gray). Sidechains are shown for the N- (S443) and C- (Y505) terminal residues of JS7 and the 4 Y residues forming hydrogen bonds across the interface. **Bottom)** Alignment showing three circulating human variants (red arrows, yellow side chains) at L452R (recent California), E484K^4^ and N501Y^15^.

We compared JS7 to four recombinant proteins obtained from other groups, including two versions of the RBD of the SARS-CoV-2. One is expressed in 293 cells (derived from human embryonic kidney cells) ^36^ and the other in yeast (proteins 1 and 4, respectively in Fig. 2), one approximately full length S protein^36^ of SARS-CoV-1 (JSP-657, protein 2), and its corresponding RBD area (RBD 219-N1^3^). The two versions of the SARS CoV-2 RBD differ in that (1) was purified from a mammalian cell line attached to a linker protein and the other (4) was purified in yeast cells ^7^ (Fig. 2). An alignment of the RBD sequences (proteins 1,3,4) is included in supplementary materials.

**Fig. 2.**
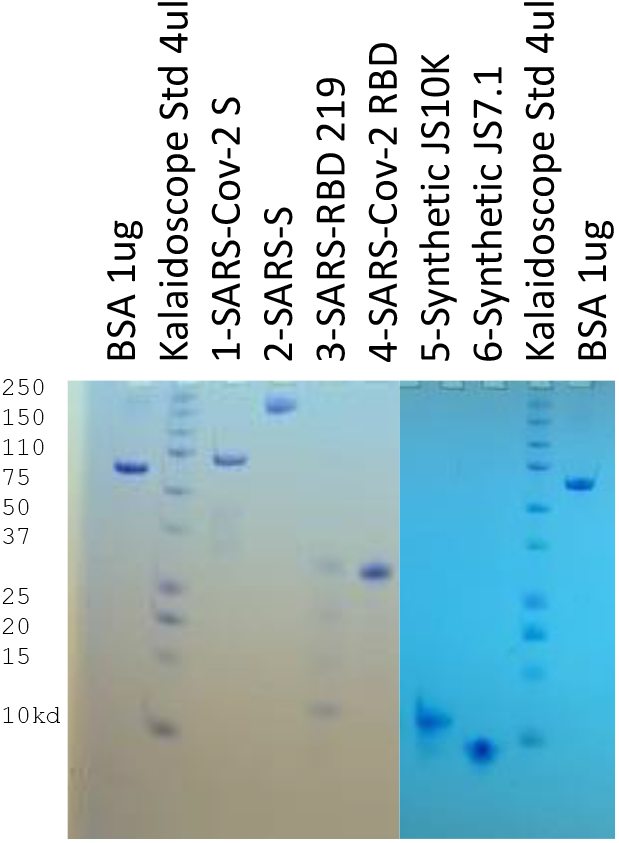
S protein and RBD fragments of SARS and SARS-CoV-2. used for dotspots and ELISA assays (je 1.25 µg). 1 &2: SARS-CoV-2 RBD-SD1 (SSM-1175)^1^ and JSP-657 SARS-Cov-1 528^2^; 3&4: SARS RBD 219-N1^3^ and SARS CoV-2 RBD^7^; 5 & 6: synthetic proteins from the ACE2 interacting area of SARS-CoV-2 (10,130D and 7158D) on 17% Tris/tricine PAGE developed with Coomassie blue gel stain.

Convalescent sera from 9 hospitalized patients with COVID-19 disease of varying severity, from “mild” to critical, were used for dotspots, ELISAs and an assay for inhibition of association of the RBD with its cellular receptor, ACE2. As shown in Table 1S, these patients had co-morbidities that have been found, in other studies, to be associated with hospitalization due to COVID-19. Of the 9, 5 were known to be diabetic, and 6 had elevated glucose levels at time of diagnosis. Four of the 6 patients who had severe or critical disease were obese (BMI>30). Notably, the one patient who succumbed did not have these risk factors except for age >70. As Fig. 3 shows, sera from patients with severe or critical disease had antibodies that interfered with the binding of the SARS-CoV-2 RBD (protein 4) to an ACE2 fragment. Sera from the two patients with mild or moderate disease did not inhibit in this assay. However, both patients had antibodies that recognized the RBD and/or the JS7 in dotspots or ELISA; this recognition was inhibited by peptides from the RBD/ACE2 interface (Fig. 4,5; see supplementary material for peptide sequences).

**Table 1.** Antibody binding to recombinant S protein and JS7 from sera of 3 COVID 19 patients and a control serum. Methods and data for all 9 patients and controls are described in Supplementary material. Recombinant proteins (1-4) or synthetic proteins (5,6) (see Fig. 2) were bound to nitrocellulose for dotspots^9^ and reacted with sera diluted 1:100. ELISA, for binding to recombinant SAR-CoV-2 RBD (Protein 4) or JS7 (6) is the highest dilution factor of serum where significant OD450 is measured; maximum dilution in the assay was 3.3 × 10^5^. ACE2 inhibition is the degree to which each serum inhibited the binding of an ACE2 fragment to SARS-CoV-2 RBD (protein 4). A negative number indicates inhibition, where protein 4 inhibition (of binding to itself) = −0.82. (average of triplicates is shown).

| Patient/age | Severity | Protein dotspot |  |  |  |  |  | Binding in ELISA to |  | ACE2 inhibition |
| --- | --- | --- | --- | --- | --- | --- | --- | --- | --- | --- |
|  |  | 1 | 2 | 3 | 4 | 5 | 6 | RBD (4) | JS7 (6) |  |
| Female/30-40 | mild | | | | | | | $4 \times 10^3$ | $3.7 \times 10^4$ | 0.26 |
| Male/50-60 | severe | | | | | | | $4 \times 10^3$ | $3.3 \times 10^5$ | -0.98 |
| Male/30-40 | critical | | | | | | | $3.3 \times 10^5$ | $3.3 \times 10^5$ | -1.28 |
| Female/20-30 | Control serum | | | | | | | $1.2 \times 10^4$ | $3.6 \times 10^4$ | -0.09 |

**Fig. 3:**
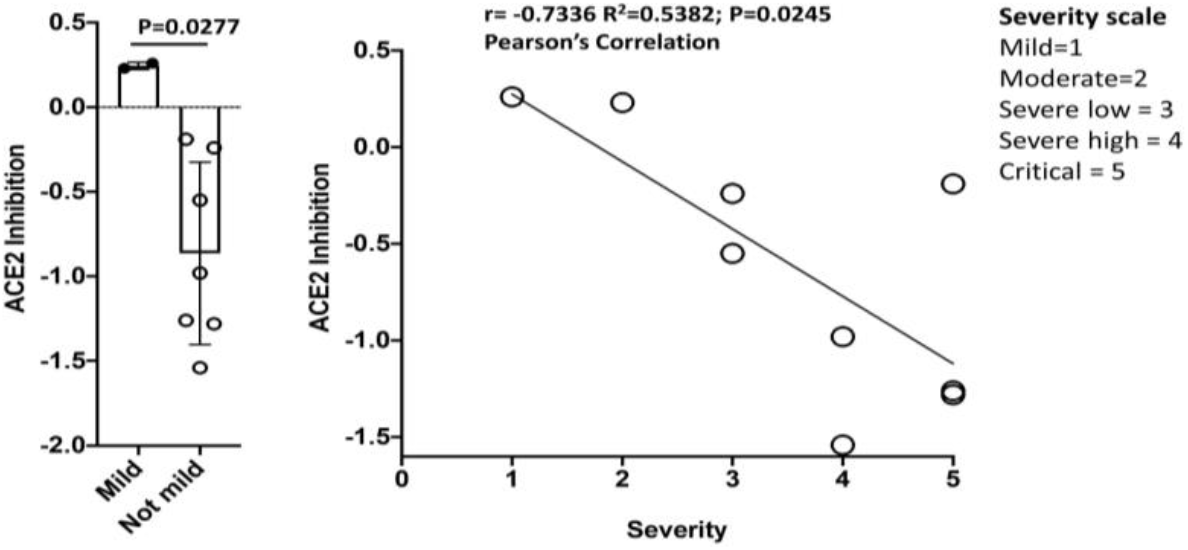
Level of antibodies in patient sera that block the interaction between the RBD (protein 4, Fig. 2) and an ACE2 receptor fragment correlate with disease severity ((decreasing OD650 indicates increasing inhibition of the interaction).

**Fig. 4:**
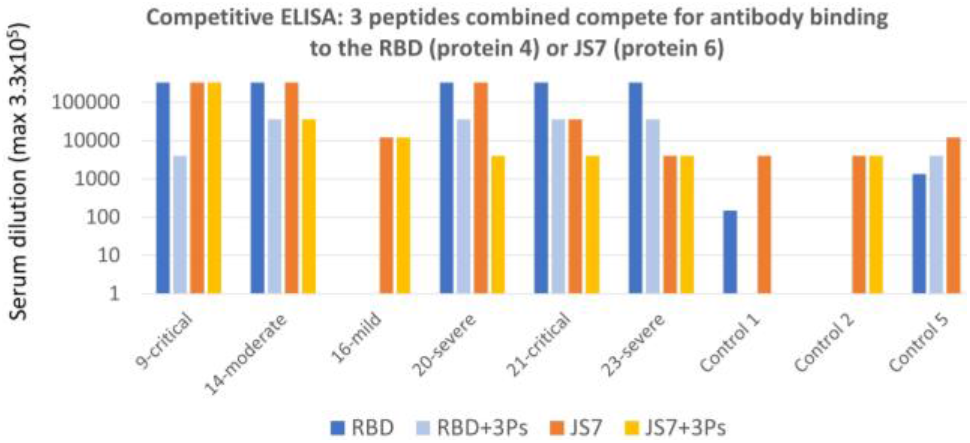
Antibody binding to the RBD of SARS-CoV-2 or JS7 (protein 4 or 6, Fig. 2, respectively) in sera of a mild, a moderate, 2 severe and 2 critical cases of COVID-19 is inhibited by 3 peptides from the RBD/ACE2 interface in competitive ELISA (maximum serum dilution to see binding). See supplementary material for peptide sequences, patient (Table S1) and control sera from spring 2019 (Table S2).

**Fig. 5:**
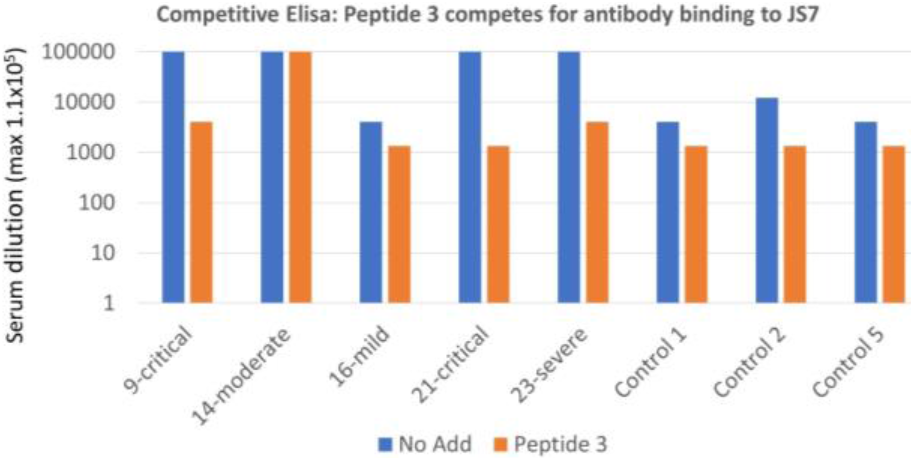
Binding to JS7 by antibodies in convalescent sera of a mild, a moderate, a severe and two critical cases of COVID-19 is inhibited by Peptide 3: SKVGGNYNYLYRLF (442-458)) from the RBD/ACE2 interface interfere in competitive ELISA (maximum serum dilution to see binding is shown).

A dotspot assay showed that all 9 patient sera contained antibodies that recognized JS7 (spot #6), while recognition of the SARS-CoV-2 recombinant RBD (Spots 1 and 4) varied (Table 1, Table S1). Binding to JS7 was similar, in ELISA assays, to that of the SARS-CoV-2 RBD (Fig. 4). While all patients had antibodies that recognized both SARS-CoV-2 RBD proteins, one patient (16) with mild disease did not recognize the full-length S protein of SARS-CoV-1 (spot 2), in two separate blood draws 5 weeks apart (Table 1).

Antibody binding, as measured by ELISA of either protein was approximately proportional to the severity of the infection (Fig. 4, 5). Antibody binding could be reduced, for some sera to the level of controls, by adding peptides from the RBD/ACE2 interface (Figure 4, 5 and Supplementary material), further emphasizing the importance of this region for binding to the cell receptor. In the case of JS7, most of the binding could be abolished by adding only one of the peptides, from the central portion of the interface with ACE2. Thus detecting evidence of prior or ongoing COVID-19 infection with the smaller synthetic protein was as sensitive as with the larger RBD, and was specific for antibodies recognizing an area essential for interacting with the ACE2. This suggests it to be an excellent candidate for point of care diagnostics that could be used, in addition to PCR, to determine immune system involvement.

Assays with sera from normal volunteers who participated in a study of chronic urticaria, collected in Denver, CO USA in spring, 2019 before widespread introductions of SARS-Cov-2 into the US (Table S2), illustrates one of the great advantages to using the small synthetic proteins for diagnostics. Larger recombinant proteins may need to be expressed with protein tags, for solubility and/or ease in purification. As the strong binding of both control and patient sera to protein 1, which still contains such tags, shows these can cause assay artifacts if left intact (Tables 1, S1, S2). Antibodies in all the control sera showed somewhat less recognition of the RBD fragment of both SARS-CoV-1 and SARS-CoV-2 produced in yeast (proteins 3,4; the sequences of all 3 forms of the RBD are compared in Supplementary Material). Two of the control sera bound three of the four recombinant proteins to the same extent as patients with only mild COVID-19 disease; this was confirmed by ELISA (Figs. 4,5). This binding could be due to previous infections with other coronaviruses^37^, or simply reflect germline antibody recognition. In support of the latter, others have found that neutralizing monoclonal antibodies against the SARS-CoV-2 S protein, isolated from many different donors, were from public clonotypes, bound autoantigens and contained relatively few somatic mutations from germline^38^.

The greatest advantage of using JS7 for diagnostic purposes is that its sequence can be rapidly modified to reflect variants now emerging in patients throughout the world. Although the N501Y mutant did not alter neutralization by polyclonal antibodies in some test sera ^39^, other changes in this area at the interface of the RBD and its ACE2 receptor (Fig. 1B) may alter the usefulness of treatments, such as convalescent plasma^4^, monoclonal antibodies and small molecules designed to disrupt this interaction.

Several lines of evidence indicate that neutralization by serum antibodies and especially monoclonal antibodies is limited by variation in the JS7 area of the SARS viruses. The affinity of the cross reactive SARS-CoV-1 antibody, CR3022, whose epitope is located in the conserved area just upstream of the JS7 site,^28^ was greatly reduced by the single amino acid change, P to A found in the SARS-CoV-2 protein.^29^ Further, while all 9 COVID-19 patients in this study had antibodies that recognize the RBD of SARS-CoV-2, one patient with mild illness did not recognize the whole S protein of SARS-CoV-1 (protein 2), despite the 80% overall sequence identity (Table 1, S1). Further, some variants in this area, which also arose as escape mutants of monoclonal antibodies^40^, can render the virus insensitive to neutralization by convalescent sera^41^. The rise of these variants in several different areas of the world suggests they do not prevent virus replication and may be selected for under immune pressure.

In this day of very specific treatments, assays which discriminate among variants of SARS related viruses and other coronaviruses are essential. It is thus important to have a panel of antigens representing areas of maximum variation in the RBD that can be used to rapidly determine specific variants and support planning clinical treatments. Reagents and linkers needed for assays can be integrated into variants of JS7 during synthesis, while controlling its sequence, stereochemistry and disulfide patterns. The JS7 protein, further modified to display variants and to be serum stable, may also have a future as a vaccine additive or booster. For example, recent methods have been developed to generate one component-synthetic proteins with incorporated adjuvant^42^.

## In conclusion

We show here that short synthetic proteins, which can be produced quickly in large quantities, can be the basis of specific assays to detect antibodies in sera that that are produced in response to infection with SARS-CoV-2. Antibodies in sera of 9 patients who had PCR confirmed, COVID-19 of differing severity recognized the JS7 protein. The sera from 8 of these patients was taken up to 2 months after diagnosis, while the 9^th^ patient died during hospitalization. JS7 and future variants should prove to be another tool for eventually defeating the current outbreak.

## Supporting information

Supplementary for Synthetic Proteins for COVID-19 Diagnostics

## Data Availability

All data is either included in the paper, supplementary material, or available from the authors

## Acknowledgements

We thank Wendy S. Baker for expert technical assistance, and Wen-Hsian Chen and others in the group at Baylor College of Medicine and Daniel Wrapp (Dartmouth) and the group of Jason McLellan of the University of Texas at Austin for supplying the recombinant proteins (1-4 in Fig 2) used throughout this work. Peptide and protein syntheses at LANL were supported by LDRD ER funding.

## Literature cited

1. Wec, A.Z. et al. Broad neutralization of SARS-related viruses by human monoclonal antibodies. Science 369, 731–736 (2020).

2. Pallesen, J. et al. Immunogenicity and structures of a rationally designed prefusion MERS-CoV spike antigen. Proceedings of the National Academy of Sciences of the United States of America 114, E7348–E7357 (2017).

3. Chen, W.H. et al. Optimization of the Production Process and Characterization of the Yeast-Expressed SARS-CoV Recombinant Receptor-Binding Domain (RBD219-N1), a SARS Vaccine Candidate. Journal of pharmaceutical sciences 106, 1961–1970 (2017).

4. Wibmer, C.K. et al. SARS-CoV-2 501Y.V2 escapes neutralization by South African COVID-19 donor plasma. bioRxiv : the preprint server for biology, 2021.2001.2018.427166 (2021).

5. Paraskevis, D. et al. Full-genome evolutionary analysis of the novel corona virus (2019-nCoV) rejects the hypothesis of emergence as a result of a recent recombination event. Infection, genetics and evolution : journal of molecular epidemiology and evolutionary genetics in infectious diseases 79, 104212 (2020).

6. Giovanetti, M., Benvenuto, D., Angeletti, S. & Ciccozzi, M. The first two cases of 2019-nCoV in Italy: Where they come from? Journal of medical virology (2020).

7. Chen, W.-H. et al. Cloning, Expression and Biophysical Characterization of a Yeast-expressed Recombinant SARS-CoV-2 Receptor Binding Domain COVID-19 Vaccine Candidate. bioRxiv : the preprint server for biology, 2020.2011.2009.373449 (2020).

8. Walls, A.C. et al. Structure, Function, and Antigenicity of the SARS-CoV-2 Spike Glycoprotein. Cell 181, 281–292 e286 (2020).

9. Baker, W.S., Negi, S., Braun, W. & Schein, C.H. Producing physicochemical property consensus alphavirus protein antigens for broad spectrum vaccine design. Antiviral research 182, 104905 (2020).

10. Walls, A.C. et al. Unexpected Receptor Functional Mimicry Elucidates Activation of Coronavirus Fusion. Cell 176, 1026–1039 e1015 (2019).

11. Pinto, D. et al. Cross-neutralization of SARS-CoV-2 by a human monoclonal SARS-CoV antibody. Nature 583, 290–295 (2020).

12. Rota, P.A. et al. Characterization of a novel coronavirus associated with severe acute respiratory syndrome. Science 300, 1394–1399 (2003).

13. Pollet, J. et al. SARS-CoV-2 RBD219-N1C1: A Yeast-Expressed SARS-CoV-2 Recombinant Receptor-Binding Domain Candidate Vaccine Stimulates Virus Neutralizing Antibodies and T-cell Immunity in Mice. bioRxiv : the preprint server for biology, 2020.2011.2004.367359 (2020).

14. Barnes, C.O. et al. Structures of Human Antibodies Bound to SARS-CoV-2 Spike Reveal Common Epitopes and Recurrent Features of Antibodies. Cell 182, 828–842 e816 (2020).

15. Wise, J. Covid-19: New coronavirus variant is identified in UK. BMJ 371, m4857 (2020).

16. Tian, X. et al. Potent binding of 2019 novel coronavirus spike protein by a SARS coronavirus-specific human monoclonal antibody. Emerging microbes & infections 9, 382–385 (2020).

17. Allam, M. et al. Genome Sequencing of a Severe Acute Respiratory Syndrome Coronavirus 2 Isolate Obtained from a South African Patient with Coronavirus Disease 2019. Microbiol Resour Announc 9 (2020).

18. Benvenuto, D. et al. The global spread of 2019-nCoV: a molecular evolutionary analysis. Pathogens and global health, 1–4 (2020).

19. Wu, C. et al. Furin: A Potential Therapeutic Target for COVID-19. iScience 23, 101642 (2020).

20. Korber, B. et al. Tracking Changes in SARS-CoV-2 Spike: Evidence that D614G Increases Infectivity of the COVID-19 Virus. Cell (2020).

21. Mansbach, R.A. et al. The SARS-CoV-2 Spike Variant D614G Favors an Open Conformational State. bioRxiv : the preprint server for biology (2020).

22. Plante, J.A. et al. Spike mutation D614G alters SARS-CoV-2 fitness. Nature (2020).

23. Liu, S. et al. Genetic Spectrum and Distinct Evolution Patterns of SARS-CoV-2. Frontiers in microbiology 11, 593548 (2020).

24. Braun, B.A., Schein, C.H. & Braun, W. D-graph clusters flaviviruses and beta-coronaviruses according to their hosts, disease type and human cell receptors. bioRxiv : the preprint server for biology (2020).

25. Lam, S.D. et al. SARS-CoV-2 spike protein predicted to form complexes with host receptor protein orthologues from a broad range of mammals. Scientific reports 10, 16471 (2020).

26. Hammer, A.S. et al. SARS-CoV-2 Transmission between Mink (Neovison vison) and Humans, Denmark. Emerging infectious diseases 27 (2020).

27. Perera, R.A. et al. Serological assays for severe acute respiratory syndrome coronavirus 2 (SARS-CoV-2), March 2020. Euro surveillance : bulletin Europeen sur les maladies transmissibles = European communicable disease bulletin 25 (2020).

28. Yuan, M. et al. A highly conserved cryptic epitope in the receptor binding domains of SARS-CoV-2 and SARS-CoV. Science 368, 630–633 (2020).

29. Wu, N.C. et al. A natural mutation between SARS-CoV-2 and SARS-CoV determines neutralization by a cross-reactive antibody. PLoS pathogens 16, e1009089 (2020).

30. Zhu, N. et al. A Novel Coronavirus from Patients with Pneumonia in China, 2019. The New England journal of medicine 382, 727–733 (2020).

31. Bowen, D.M., Lewis, J.A., Lu, W. & Schein, C.H. Simplifying complex sequence information: A PCP-consensus protein binds antibodies against all four Dengue serotypes. Vaccine 30, 6081–6087 (2012).

32. Schein, C.H. et al. Physicochemical property consensus sequences for functional analysis, design of multivalent antigens and targeted antivirals. BMC Bioinformatics 13 Suppl 13, S9 (2012).

33. Danecek, P., Lu, W. & Schein, C.H. PCP consensus sequences of flaviviruses: correlating variance with vector competence and disease phenotype. J Mol B iol 396, 550–563 (2010).

34. Yuan, M., Liu, H., Wu, N.C. & Wilson, I.A. Recognition of the SARS-CoV-2 receptor binding domain by neutralizing antibodies. Biochemical and b iophysical research communications (2020).

35. Zhou, T. et al. Cryo-EM Structures of SARS-CoV-2 Spike without and with ACE2 Reveal a pH-Dependent Switch to Mediate Endosomal Positioning of Receptor-Binding Domains. Cell host & microbe 28, 867–879 e865 (2020).

36. Wrapp, D. et al. Cryo-EM structure of the 2019-nCoV spike in the prefusion conformation. Science 367, 1260–1263 (2020).

37. Ladner, J.T. et al. Epitope-resolved profiling of the SARS-CoV-2 antibody response identifies cross-reactivity with endemic human coronaviruses. Cell Reports Medicine 2, 100189 (2021).

38. Kreye, J. et al. A Therapeutic Non-self-reactive SARS-CoV-2 Antibody Protects from Lung Pathology in a COVID-19 Hamster Model. Cell 183, 1058–1069 e1019 (2020).

39. Xie, X. et al. Neutralization of N501Y mutant SARS-CoV-2 by BNT162b2 vaccine-elicited sera. bioRxiv : the preprint server for biology, 2021.2001.2007.425740 (2021).

40. Liu, Z. et al. Landscape analysis o f escape variants identifies SARS-CoV-2 spike mutations that attenuate monoclonal and serum antibody neutralization. bioRxiv : the preprint server for biology (2020).

41. Andreano, E. et al. SARS-CoV-2 escape in vitro from a highly neutralizing COVID-19 convalescent plasma. bioRxiv : the preprint server for biology (2020).

42. Hanna, C.C. et al. Synthetic protein conjugate vaccines provide protection against Mycobacterium tuberculosis in mice. Proceedings of the National Academy of Sciences 118, e2013730118 (2021).

