## Supplementary for Synthetic Proteins for COVID-19 Diagnostics for "Synthetic protein antigens for COVID-19 diagnostics"

**Figure S1A**: alignment of the SARS-CoV-1 (SARS2003) and SARS-CoV-2 sequences (Wuhan China, January, 2020) with the regions of maximum variability and the Furin cleavage insert in the latter virus[1] highlighted in yellow. The longest region of variability in the ACE2 binding region was chosen for synthesis. The ~ 300 C-terminal residues (not shown) are remarkably conserved between the two viruses. The positions of three changes (deletion 69-70, N501Y, P to H at the Furin cleavage insertion) in the recent circulating strain VUI-202012/01 (lineage B.1.1.7) are marked with arrows.

```
CLUSTAL O(1.2.4) multiple sequence alignment


SARS2003CDC          MFIFLLFLTLTSGSDLDRCTTFDDVQAPNYTQHTSSMRGVYYPDEIFRSDTLYLTQDLFL     60
SARS_CoV_2CHN2020    MFVFLVLLPLVSSQCVNLTT--RTQLPPAY--TNSFTRGVYYPDKVFRSSVLHSTQDLFL     56
                     **:**::* *.*.. ::  *       * *   .*  *******::***..*: ******

SARS2003CDC          PFYSNVTGFHTIN-------HTFGNPVIPFKDGIYFAATEKSNVVRGWVFGSTMNNKSQS     113
SARS_CoV_2CHN2020    PFFSNVTWFHAIHVSGTNGTKRFDNPVLPFNDGVYFASTEKSNIIRGWIFGTTLDSKTQS     116
                     **:**** **:*:       : *.***:**:**:***:*****::***:**:*::.*:**

SARS2003CDC          VIIINNSTNVVIRACNFELCDNPFFAVSKPMGT----QTHTMIFDNAFNCTFEYISDAFS     169
SARS_CoV_2CHN2020    LLIVNNATNVVIKVCEFQFCNDPFLGVYYHKNNKSWMESEFRVYSSANNCTFEYVSQPFL     176
                     ::*:**:*****:.*:*::*::**:.*    ..    ::.  ::..* ******:*: *

SARS2003CDC          LDVSEKSGNFKHLREFVFKNKDGFLYVYKGYQPIDVVRDLPSGFNTLKPIFKLPLGINIT     229
SARS_CoV_2CHN2020    MDLEGKQGNFKNLREFVFKNIDGYFKIYSKHTPINLVRDLPQGFSALEPLVDLPIGINIT     236
                     :*:. *.****:******** **:: :*. : **::*****.**.:*:*:..**:*****

SARS2003CDC          NFRAILTAFS------PAQDIWGTSAAAYFVGYLKPTTFMLKYDENGTITDAVDCSQNPL     283
SARS_CoV_2CHN2020    RFQTLLALHRSYLTPGDSSSGWTAGAAAYYVGYLQPRTFLLKYNENGTITDAVDCALDPL     296
                     .*:::*: .       :.. * :.****:****:* **:***:***********: :**

SARS2003CDC          AELKCSVKSFEIDKGIYQTSNFRVVPSGDVVRFPNITNLCPFGEVFNATKFPSVYAWERK     343
SARS_CoV_2CHN2020    SETKCTLKSFTVEKGIYQTSNFRVQPTESIVRFPNITNLCPFGEVFNATRFASVYAWNRK     356
                     :* **::*** ::*********** *: .:******************:* *****:**

SARS2003CDC          KISNCVADYSVLYNSTFFSTFKCYGVSATKLNDLCFSNVYADSFVVKGDDVRQIAPGQTG     403
SARS_CoV_2CHN2020    RISNCVADYSVLYNSASFSTFKCYGVSPTKLNDLCFTNVYADSFVIRGDEVRQIAPGQTG     416
                     :**************: ********** ********:********::**:**********

SARS2003CDC          VIADYNYKLPDDFMGCVLAWNTRNIDATSTGNYNYKYRYLRHGKLRPFERDISNVPFSPD     463
SARS_CoV_2CHN2020    KIADYNYKLPDDFTGCVIAWNSNNLDSKVGGNYNYLYRLFRKSNLKPFERDISTEIYQAG     476
                      ************ ***:***:.*:*:.   ***** ** :*:.:*:*******.  :. .

SARS2003CDC          GKPCTP-PALNCYWPLNDYGFYTTTGIGYQPYRVVVLSFELLNAPATVCGPKLSTDLIKN     522
SARS_CoV_2CHN2020    STPCNGVEGFNCYFPLQSYGFQPTNGVGYQPYRVVVLSFELLHAPATVCGPKKSTNLVKN     536
                     ..**.    .:***:**:.***  *.*:***************:********* **:*:**

SARS2003CDC          QCVNFNFNGLTGTGVLTPSSKRFQPFQQFGRDVSDFTDSVRDPKTSEILDISPCSFGGVS     582
SARS_CoV_2CHN2020    KCVNFNFNGLTGTGVLTESNKKFLPFQQFGRDIADTTDAVRDPQTLEILDITPCSFGGVS     596
                     :*************** *.*:* ********::* **:****:* *****:********

SARS2003CDC          VITPGTNASSEVAVLYQDVNCTDVSTAIHADQLTPAWRIYSTGNNVFQTQAGCLIGAEHV     642
SARS_CoV_2CHN2020    VITPGTNTSNQVAVLYQDVNCTEVPVAIHADQLTPTWRVYSTGSNVFQTRAGCLIGAEHV     656
                     *******:*.:***********:* .*********:**:****.*****:**********

SARS2003CDC          DTSYECDIPVGAGICASYHTVSL----LRSTSQKSIVAYTMSLGADSSIAYSNNTIAIPT     698
SARS_CoV_2CHN2020    NNSYECDIPIGAGICASYQTQTNSPRRARSVASQSIIAYTMSLGAENSVAYSNNSIAIPT     716
                     :.*******:********:* :      **.:.:**:********:.*:*****:*****

SARS2003CDC          NFSISITTEVMPVSMAKTSVDCNMYICGDSTECANLLLQYGSFCTQLNRALSGIAAEQDR     758
SARS_CoV_2CHN2020    NFTISVTTEILPVSMTKTSVDCTMYICGDSTECSNLLLQYGSFCTQLNRALTGIAVEQDK     776
                     **:**:***::****:******.**********:*****************:***.***:

SARS2003CDC          NTREVFAQVKQMYKTPTLKYFGGFNFSQILPDPLKPTKRSFIEDLLFNKVTLADAGFMKQ     818
SARS_CoV_2CHN2020    NTQEVFAQVKQIYKTPPIKDFGGFNFSQILPDPSKPSKRSFIEDLLFNKVTLADAGFIKQ     836
                     **:********:****  :* ************* **:*********************:**

SARS2003CDC          YGECLGDINARDLICAQKFNGLTVLPPLLTDDMIAAYTAALVSGTATAGWTFGAGAALQI     878
SARS_CoV_2CHN2020    YGDCLGDIAARDLICAQKFNGLTVLPPLLTDEMIAQYTSALLAGTITSGWTFGAGAALQI     896
                     **:***** **********************:*** **:**::** *:************

SARS2003CDC          PFAMQMAYRFNGIGVTQNVLYENQKQIANQFNKAISQIQESLTTTSTALGKLQDVVNQNA     938
SARS_CoV_2CHN2020    PFAMQMAYRFNGIGVTQNVLYENQKLIANQFNSAIGKIQDSLSSTASALGKLQDVVNQNA     956
                     ************************* ******.**.:**:**::*::************
```

**Fig. S1B: Alignment of the SARS-CoV-2 S protein sequence from Wuhan China, January 2020 with SARS-CoV-1 (protein 3) [3] and SARS-CoV-2 RBDs (Proteins 1[1] and 4[14]) used in this study, numbered according to their order on the protein gel of Figure 2 and the dotspots shown throughout this work.** The JS7 sequence (protein 6) is highlighted in yellow. Proteins 3 and 4 were expressed in yeast[14]. Protein 1 was expressed in FreeStyle 293 cells, which are derived from human embryonic kidney cells; the sequence below (residues 319-591) was followed by a C-terminal HRV3C protease cleavage site, a monomeric Fc tag and an 8XHisTag[1].

```
SARS_CoV_2CHN2020      CTLKSFTVEKGIYQTSNFRVQPTESIVRFPNITNLCPFGEVFNATRFASVYAWNRKRISN  360
JSM1175-SARSCoV2RBD(1) ------------------RVQPTESIVRFPNITNLCPFGEVFNATRFASVYAWNRKRISN  42
SARS-CoV-1 RBD(3)      -------------------------------ITNLCPFGEVFNATKFPSVYAWERKKISN  29
SARS-CoV-2-RBD (4)     -------------------------------ITNLCPFGEVFNATRFASVYAWNRKRISN  29
                                                      **************:* *****:**:***

SARS_CoV_2CHN2020      CVADYSVLYNSASFSTFKCYGVSPTKLNDLCFTNVYADSFVIRGDEVRQIAPGQTGKIAD  420
JSM-1175 (1)           CVADYSVLYNSASFSTFKCYGVSPTKLNDLCFTNVYADSFVIRGDEVRQIAPGQTGKIAD  102
SARS-CoV (3)           CVADYSVLYNSTFFSTFKCYGVSATKLNDLCFSNVYADSFVVKGDDVRQIAPGQTGVIAD  89
SARS-CoV-2(4)          CVADYSVLYNSASFSTFKCYGVSPTKLNDLCFTNVYADSFVIRGDEVRQIAPGQTGKIAD  89
                       ***********: ********** ********:********::**:********** ***

SARS_CoV_2CHN2020      YNYKLPDDFTGCVIAWNSNNLDSKVGGNYNYLYRLFRKSNLKPFERDISTEIYQAGSTPC  480
JSM-1175 (1)           YNYKLPDDFTGCVIAWNSNNLDSKVGGNYNYLYRLFRKSNLKPFERDISTEIYQAGSTPC  162
SARS-CoV (3)           YNYKLPDDFMGCVLAWNTRNIDATSTGNYNYKYRYLRHGKLRPFERDISNVPFSPDGKPC  149
SARS-CoV-2(4)          YNYKLPDDFTGCVIAWNSNNLDSKVGGNYNYLYRLFRKSNLKPFERDISTEIYQAGSTPC  149
                       ********* ***:***:.*:*:.  ***** ** :*:.:*:*******.  :. ...**

SARS_CoV_2CHN2020      NGVEGFNCYFPLQSYGFQPTNGVGYQPYRVVVLSFELLHAPATVCGPKKSTNLVKNKCVN  540
JSM-1175 (1)           NGVEGFNCYFPLQSYGFQPTNGVGYQPYRVVVLSFELLHAPATVCGPKKSTNLVKNKCVN  222
SARS-CoV (3)           TP-PALNCYWPLNDYGFYTTTGIGYQPYRVVVLSFELLNAPATVCGPKLSTDLIKNQCVN  208
SARS-CoV-2(4)          NGVEGFNCYFPLQSYGFQPTNGVGYQPYRVVVLSFELLHAPATVCGPKKSTNLVKNKCVN  209
                       .   .:***:**:.***  *.*:***************:********* **:*:**:***

SARS_CoV_2CHN2020      FNFNGLTGTGVLTESNKKFLPFQQFGRDIADTTDAVRDPQTLEILDITPCSFGGVSVITP  600
JSM-1175  (1)          FNFNGLTGTGVLTESNKKFLPFQQFGRDIADTTDAVRDPQTLEILDITPCSGSLE-VLFQ  281
SARS-CoV (3)           FNFNGLTGTG--------------------------------------------------  218
SARS-CoV-2 (4)         FNFNGLTGTHHHHHH---------------------------------------------  224
                       *********
```

**Figure S2:** Characterizing the JS7 protein.

**A)** The protein was purified with HPLC to homogeneity (left). The MS-ESI (right) indicates, consistent with sequence, an average molecular mass to be 7157.9.

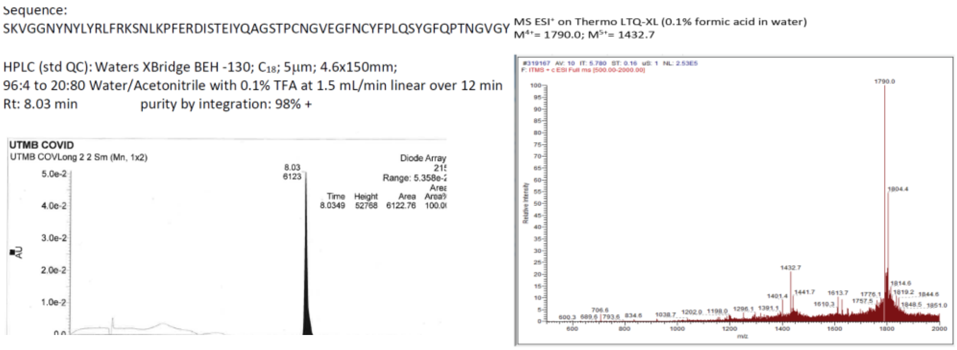


**B)** Left: CD spectrum overlaid with the calculated result (NRMSD:0.006) from the CDSSTR program indicates some β-strand content, consistent with the experimental structure of the S protein (right). JS7

was diluted to ~0.2 mg/ml in water. The method used reference data set: 1  (Sreerama, N. and Woody, R.W. (2000), Analytical Biochemistry, 287, 252–260 and references therein)

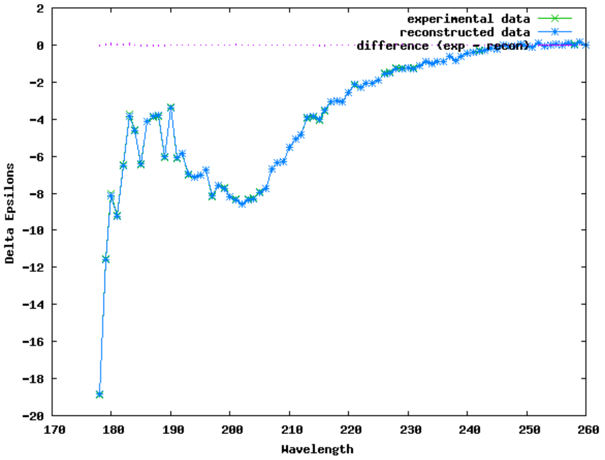


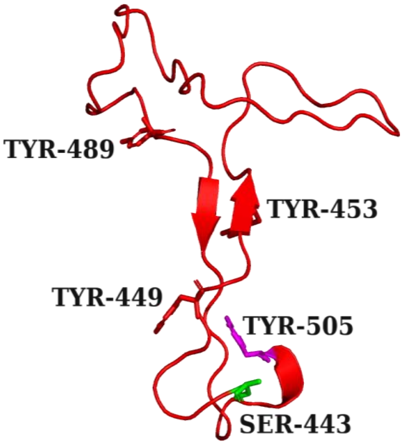


**Calculated Secondary Structure Fractions**

| Helix1 | Helix2 | Strand1 | Strand2 | Turns | Unordered | Total |
|---|---|---|---|---|---|---|
| 0.00 | -.01 | 0.37 | 0.21 | 0.27 | 0.18 | 1.02 |

**Table S1**: Clinical details, treatment outcome and assay results for 9 patients admitted for PCR-confirmed, Covid-19 related pneumonia, whose sera were used in this study. Abbreviations indicating the course of disease: LOS/ICU LOS: length of stay in days; ACE2 inhibition: as measured as described below;  SD: standard deviation (of 3 samples). Data for Diabetes (Type 2), Blood glucose (in mg/dl, normal range is 70-110) are at admission. All samples were diluted 1:100 for the dotspots, except the Invitrogen rabbit serum, which was 1:200. See Materials and Methods for assay details. Note that patient 0016 sera was similar to the control sera (Table 2S) in that it did not recognize the SARS-CoV-1 S protein (spot 2) in either of two samples taken one and two months after PCR negative status for the virus. Sera from patients with severe disease prevented binding of a fragment of the ACE2 cell receptor to the SARS-CoV-2 RBD (protein 4), although those with mild illness did not (Fig. 3 in the text). Adding the SARS-CoV-2 protein in PBS to this assay gave an average inhibition (decrease in $OD_{450}$ compared to buffer alone) of -0.82.

| Record ID | Sex | Age | O2 therapy | Outcome | Clinical Pneumonia | Diabetes | Glucose | Severity* | Dotspot | ACE2 inhibition | SD |
|---|---|---|---|---|---|---|---|---|---|---|---|
| OBS-0006 | F | 46-70 | None | Discharged alive | Yes | Yes | 80-125 | Moderate-2 | 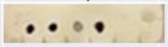 | 0.23 | 0.03 |
| OBS-0009 | M | 20-45 | Invasive Vent | Home Health | Yes | Yes | High | Critical-5 | 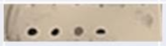 | -0.19 | 0.06 |
| OBS-0014 | M | 46-70 | Nasal Canula | Discharged alive | Yes | No | 80-125 | Severe low-3 | 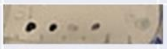 | -0.24 | 0.09 |
| OBS-0015 | M | 20-45 | Invasive Vent | Transferred to LTAC | Yes | Yes | High | Critical-5 | 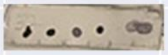 | -1.28 | 0.17 |
| OBS-0016 | F | 20-45 | None | Discharged alive | No | No | 80-125 | Mild-1 | 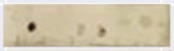 | 0.26 | 0.02 |
| OBS-0018 | M | 20-45 | Nasal Canula | Discharged alive | Yes | No | 80-125 | Severe low-3 | 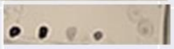 | -0.55 | 1.07 |
| OBS-0020 | M | 46-70 | Hi-flow Nasal Canula | Discharged alive | Yes | Yes | High | Severe high-4 | 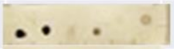 | -0.98 | 0.04 |
| OBS-0021 | F | >70 | Invasive Vent | Death | Yes | No | 80-125 | Critical-5 | 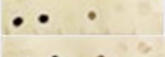 | -1.26 | 0.29 |
| OBS-0023 | F | 46-70 | BiPAP | Discharged alive | Yes | Yes | High | Severe high-4 | 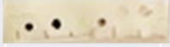 | -1.54 | 0.17 |
| | | | | | | | Protein dotted | | 1 2 3 4 5 6 | | |
| | | | | | | Rabbit serum Invitrogen 1:200 | | | 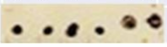 | | |

*Severity score assigned according to:
https://www.covid19treatmentguidelines.nih.gov/overview/clinical-spectrum/
1. Mild (symptoms, but not lower respiratory symptoms)
2. Moderate (some evidence of lower respiratory, but no $O_2$ needs)
3. Severe – low (needed $O_2$, but not much)
4. Severe – high (needed high delivery $O_2$ device)
5. Critical – intubated

**Table S2: Control samples collected in Spring, 2019 in Denver CO:** These are control sera from a study of chronic urticaria, approved by local IRB; all participants (ages 24-30, sera were collected in Feb-April, 2019) gave informed consent for sera to be used in immunology research. Sera were diluted 1:20 and then 1:100 depending on the strength of the spots after the first dilution. The RBD protein from SARS-CoV-2, which contains a fusion protease tag (spot 1) is recognized by all of the control sera, but the full length S protein of SARS-CoV-1 (spot 2) is not. The RBD of both SARS-CoV-1 (spot 3) and SARS-CoV-2 (spot 4) purified from yeast are also recognized by two of the controls, while the two synthetic proteins (JS 10 and 7, Spots 5 and 6) are only slightly recognized.

| Code | Gender | Race | Dotspot (1:20, 1:100) Protein: 1 2 3 4 5 6 | |
|---|---|---|---|---|
| Control 1 | M | Caucasian | 1:20 | 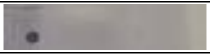 |
| Control 2 | F | Asian | 1:20 | 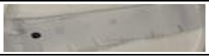 |
| Control 3 | F | Caucasian | 1:100 | 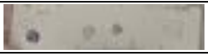 |
| Control 4 | M | Other | 1:100 | 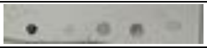 |
| Control 5 | F | Caucasian | 1:100 | 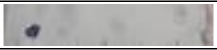 |

**Materials and Methods**

Sera: were collected under separate protocols, are de-identified discarded samples, and are thus considered exempt. Table S1 summarizes the patient characteristics, treatment, and disease outcomes.

Human sera after infection with COVID-19 ( 10 samples from 9 patients (Table S1)) were negative for residual virus presence. De-identified clinical samples and clinical data were collected from consented patients under the Observational Protocol for Diseases and Exposures of Public Health Importance (UNMC IRB # 060-20-EP/UTMB-IRB # 20-0031), PI, Dr. Mark Kortepeter, U. Nebraska Medical Center; UTMB site-PI, Dr. Susan McLellan, developed by the Special Pathogens Research Network (SPRN) of the National Emerging and Special Pathogens Training and Education Center (NETEC). NETEC and SPRN are funded by the US Department of Health and Human Services Office of the Assistant Secretary for Preparedness and Response (ASPR), CFDA #93.825.

1) Five control sera (from normal individuals) were collected in the US in February-April of 2019 (Table S2) with informed consent under Colorado Multiple Institution Review Board (COMIRB) 00-802, "Redefining the Major Peanut Allergens". All samples are de-identified.

Proteins: Full length recombinant S proteins from SARS and SARS-Cov-2 were obtained from Daniel Wrapp (Dartmouth College) and purified as described [2]. Recombinant S protein fragments of the receptor binding domain (RBD), purified from yeast, were received from Wen- Hsiang Chen, Baylor College of Medicine[3,4]. Peptides and synthetic proteins (defined as >35 amino acids) from the RBD were synthesized in the Peptide Core at Los Alamos National Labs.

Peptide and Protein Synthesis: All reagents and solvents deployed were of peptide synthesis or biotech grade. All amino acids were purchased from P3Bio with the exception of Fmoc His (tBoc), required for high temperature coupling reactions which was obtained from CEM. Dimethylformamide (DMF) and the deployed deprotection reagent, 20% Pyrrole solution in DMF were obtained from Alfa Aesar. The peptide coupling reagents, Diisopropylcarbodiimide (DIC) and Oxyma Pure (Eythylcyanohyroxyiminoacetate) were acquired in peptide synthesis grade from AKScientific. General reagents such as N,N-diisopropyl ethyl amine (DIPEA), triisopropyl silane (isoPr3SiH; TIPS), thioanisole, octaethylenglycol-dithiol and trifluoroacetic acid (TFA) were purchased from Sigma Aldrich and methylene chloride (DCM) was obtained from Fisher Scientific. For HPLC purifications, Acetonitrile was acquired from Alfa Aesar and water was purified in-house (deionized, filtered through a Nanopure to 18.2 MΩ*cm resistivity, and UV-sterilized). Mass spectrometry used highest quality (Optima MS grade) solvents purchased from Fisher Scientific.

Automated Peptide Synthesis using the CEM Liberty Prime Microwave Peptide Synthesizer. A CEM Liberty Prime microwave peptide synthesizer was used for solid phase synthesis at high temperature ($105^0$C). All syntheses were performed at the 0.1 mM scale at the recommended standard instrument chemistry on a Rink amide resin. For the shorter peptides in this publication, single coupling instrument cycles were used, with achieved average coupling yields for cycle of 98.5%. For JS7 and JS10 the reaction cycles, double coupling of the amino acids was used and the drain times were increased from 5 to 10 sec to accommodate for the resin volume increase over the synthesis cycles. To prevent hydrolysis of acid labile side chain protecting groups during the extended sequence syntheses, 0.1 M DIPEA was added to the Oxyma solution. Average coupling yields thus achieved exceed 99% and even the longest sequences were obtained in moderate yield.

Deprotection and Removal of the Peptides and Proteins from the Resin: Deprotection used 25 mL of modified "reagent K" mixture: TIPS (1.25 ml/25 ml), thioanisole (0.625 ml/25 ml), octaethyleneglycoldithiol (1.25 mL/25mL), a less odorous substitute for EDT (ethylendithiol), and water (1.25 ml/25 ml) in TFA (trifluoroacetic acid). The resin was pretreated with the quencher solution for 5 min, then TFA was added (to final volume of 25 ml). The deprotections were caried out in 50 mL conical tubes of high-density PP under a blanket of Argon to prevent side reactions from air. The solutions were filtered and the filtrate was concentrated to 10 mL. The peptide was then precipitated into ice cold ether and collected by centrifugation.

Purification and Analysis of the Peptides: Purifications (to > 98% +) were performed on a Waters HPLC preparative workstation with 2545 pump (at 20 ml/min) and using a C18 reverse phase column (Waters BEH 130, 5 µm, 19x150) and a gradient from 98% to 50 % water-acetonitrile with 0.1% TFA. Peaks were collected based on monitoring at 215 nm using a PDA 2998 detector. Combined product fractions were lyophilized, yielding a white fluffy solid. Peptides were then analyzed for purity by analytical HPLC on a C18 reverse phase column (Waters BEH 130, 5 µm, 4.6x150) with a gradient from 98% to 20 % water-acetonitrile with 0.1% TFA and by mass spectrometry on Thermo LTQ or Exactive mass spectrometers, respectively, in ESI+ mode.

Sequences: JS7 (442-505 of SARS-CoV-2 S protein):

SKVGGNYNYLYRLFRKSNLKPFERDISTEIYQAGSTPCNGVEGFNCYFPLQSYGFQPTNGVGY

Peptide 1: EGF**N**CYFPLQ**S**YGFQPTNGVGY

Peptide 2: FERDISTEIYQAGST

Peptide 3: SKVGGNYNY**L**YRLF (442-458)

The bold residues in peptide 1 show two residues that when changed in SARS-CoV-1 affect ACE2 binding (N479K, T487S)[5]. These residues are most in contact with the ACE2 receptor in structures of the complex (taken from Walls et al.[6]). The bold residue **L** in Peptide 3, the most active peptide in preventing binding to JS7 (Fig. 5), is R in the “California” variant of SARS-CoV-2.

Other reagents: Positive control rabbit serum (polyclonal, against SARS/SARS-CoV-2 Coronavirus spike protein subunit 1) was Invitrogen PA5-81795.

Dotspots: Were done as described previously[7]. Proteins (0.25 µg/1 µl spot) were dotted onto nitrocellulose (Millipore, 0.2 µ) and allowed to dry. They were blocked with 5% milk in PBS, washed with PBS and incubated with diluted sera (1:100 in PBS) for 1 h at RT. The sera were removed, the dotspots washed 3 times with PBS buffer, and then incubated for 1 h in Goat-anti-human-IgG-HRP (Catalog # 2040-05) or Goat-anti-rabbit IgG-HRP (# 4050-05) from Southern Biotech, diluted 1:1000/1:2000 in PBS. After washing 3x with PBS, the spots were developed with 4-Chloronaphthol reagent.

ELISA and Competitive ELISA: Proteins were dissolved to 2 µg/ml in Borate buffer and allowed to bind to 96 well flat bottom plates (Thermofisher) overnight at 4˚C. The peptide or protein solution was removed and plates were blocked with 5% dry milk powder in PBS. The solution was discarded and the plates washed with PBS/0.1% Tween 20. Sera were assayed alone, or after mixing with the indicated concentrations of proteins or peptides, and incubated 1h at RT. In each case, they were diluted into the first row and then dilutions were made 1:3 with a multichannel pipettor down the plate. After overnight incubation at 4˚C, the serum dilutions were removed, the plates washed as above, and HRP-labeled secondary antibody (diluted 1:2000 to 1:4000 in PBS) was added. After 1 h, the plates were washed and developed with TMB reagent, using ½ volume of 2M Sulfuric acid to stop the reaction.

Assay for ACE2 binding to S protein: ACE2 receptor binding was measured with a SARS-CoV-2 inhibitor screening kit from Acrobiosystems (Catalog # EP-105), whereby the coating protein used in the assay was 2 µg/ml SARS-CoV-2 RBD (protein 4 in Fig. 2) or the JS7 synthetic protein (6 in Fig. 2). After coating with either the RBD fragment or JS7, plates were washed and blocked with 2% BSA in PBS/0.05% Tween 20. After washing, sera or other samples were applied and the plates incubated overnight at 4˚C. The plates were washed with PBS/0.05% Tween 20 and a solution of 12 µg/ml biotin conjugated ACE2 fragment was added to all wells. After incubation for 1h at 37C, the plates were washed and streptavidin coupled to HRP was added. After another 1h at 37C, the plates were washed and developed with TMB reagent and read at 652 nm before and 450 nm after addition of stop solution (1/2 volume $H_2SO_4$).


1. Wu C, Zheng M, Yang Y, et al. Furin: A Potential Therapeutic Target for COVID-19. *iScience.* 2020;23(10):101642.
2. Wrapp D, Wang N, Corbett KS, et al. Cryo-EM structure of the 2019-nCoV spike in the prefusion conformation. *Science.* 2020;367(6483):1260-1263.
3. Chen WH, Chag SM, Poongavanam MV, et al. Optimization of the Production Process and Characterization of the Yeast-Expressed SARS-CoV Recombinant Receptor-Binding Domain (RBD219-N1), a SARS Vaccine Candidate. *Journal of pharmaceutical sciences.* 2017;106(8):1961-1970.
4. Chen W-H, Wei J, Kundu RT, et al. Cloning, Expression and Biophysical Characterization of a Yeast-expressed Recombinant SARS-CoV-2 Receptor Binding Domain COVID-19 Vaccine Candidate. *bioRxiv : the preprint server for biology.* 2020:2020.2011.2009.373449.
5. Li W, Zhang C, Sui J, et al. Receptor and viral determinants of SARS-coronavirus adaptation to human ACE2. *The EMBO journal.* 2005;24(8):1634-1643.
6. Walls AC, Park YJ, Tortorici MA, Wall A, McGuire AT, Veesler D. Structure, Function, and Antigenicity of the SARS-CoV-2 Spike Glycoprotein. *Cell.* 2020;181(2):281-292 e286.
7. Baker WS, Negi S, Braun W, Schein CH. Producing physicochemical property consensus alphavirus protein antigens for broad spectrum vaccine design. *Antiviral research.* 2020;182:104905.
